# Preventable deaths from SARS-CoV-2 in England and Wales: a systematic case series of coroners’ reports during the COVID-19 pandemic

**DOI:** 10.1101/2021.07.15.21260589

**Authors:** Bethan Swift, Carl Heneghan, Jeffrey K. Aronson, David J. Howard, Georgia C. Richards

## Abstract

**Objectives:** To examine coroners’ Prevention of Future Deaths reports (PFDs) to identify deaths involving SARS-CoV-2 that coroners deemed preventable.

**Design:** Consecutive case series.

**Setting:** England and Wales.

**Participants:** Patients reported in 510 PFDs dated between 01 January 2020 and 28 June 2021, collected from the UK’s Courts and Tribunals Judiciary website using web scraping to create an openly available database, https://preventabledeathstracker.net/.

**Main outcome measures:** Concerns reported by coroners.

**Public and Patient Involvement:** Patients and members of the public were not involved in this study.

**Results:** SARS-CoV-2 was involved in 23 deaths reported by coroners in PFDs. Twelve deaths were indirectly related to the COVID-19 pandemic, defined as those that were not medically caused by SARS-CoV-2, but were associated with mitigation measures. In 11 cases the coroner explicitly reported that COVID-19 had directly caused death. There was geographical variation in the reporting of PFDs; most (39%) were written by coroners in the North-West of England. The coroners raised 56 concerns, problems in communication being the most common (30%), followed by failure to follow protocols (23%). Organizations in the National Health Service (NHS) were sent the most PFDs (51%), followed by the Government (26%), but responses to PFDs by these organizations were poor.

**Conclusions:** PFDs contain a rich source of information on preventable deaths that has previously been difficult to examine systematically. Our openly available tool (https://preventabledeathstracker.net/) streamlines this process and has identified many concerns raised by coroners that should be addressed during the Government’s inquiry into the handling of the COVID-19 pandemic, so that mistakes made are less likely to be repeated.

**Study protocol pre-registration:** https://osf.io/bfypc/

**Summary box:** *What is already known about this subject?:*

- The UK Government has stated that there will be a public inquiry into the handling of the COVID-19 pandemic, to learn lessons for future pandemics.
- Coroners in England and Wales have a duty to report and communicate information about the deaths they investigate when the coroner believes that action should be taken to prevent future deaths.
- These reports, called Prevention of Future Death reports (PFDs), had not yet been systematically analysed to identify deaths that occurred during the COVID-19 pandemic.

*What are the new findings?:*

- We created the Preventable Deaths Database (https://preventabledeathstracker.net/) using web scraping to systematically assess PFDs published on the Courts and Tribunal Judiciary website.
- Between 01 January 2020 and 28 June 2021, one in 20 (4.5%, n=23) PFDs that were published by coroners involved SARS-CoV-2.
- Coroners raised many concerns about the care of patients in hospitals, care homes, and people in the community during the COVID-19 pandemic, which require action to prevent future deaths.

*How might it affect clinical practice in the foreseeable future?:*

- Preventable deaths that occurred during the COVID-19 pandemic should be referred to the coroner so that an inquest can be performed and a PFD issued, highlightng actions that could be avoided in improving the handling of future pandemics in both the UK and elsewhere.
- Our tool, https://preventabledeathstracker.net/, can be used by others to examine preventable deaths in England and Wales, and to identify signals for quality improvement to reduce avoidable harms in clinical practice.

## Introduction

Over five million deaths worldwide have been attributed to the severe acute respiratory syndrome due to coronavirus 2 (SARS-CoV-2)^1^; some deaths may have been preventable.

In England and Wales, causes of deaths are investigated by coroners during an inquest, unless the death is natural or referred to the criminal court. Under UK regulations, coroners have a duty to report and communicate information about the deaths that they investigate when they believe that actions should be taken to prevent similar deaths^2 3,4^. Such reports, previously called Rule 43 reports, are named Prevention of Future Deaths reports or PFDs. Despite these regulations, there is no formal system in place for auditing or systematically analysing PFDs, so concerns raised by coroners in such reports may go unrecognized and unreported, reducing the impact of the PFD system.

The PFD system has three processes: 1) coroners write PFDs after the inquest and send reports to those with the power to act; 2) addressees respond to coroners regarding the concerns raised in PFDs within 56 days; and 3) actions to prevent such deaths are proposed and ideally implemented. In December 2020, a series was launched in BMJ Evidence Based Medicine to disseminate PFDs that highlight lessons for clinical practice and policy^5^. Case reports in the series have identified deaths from ingesting alcohol-based hand sanitizer, misdiagnosed adverse drug reactions, problems with repeat opioid prescriptions, and fire hazards from emollient products^6-9.^ Case series of PFDs have also been conducted to investigate preventable deaths from medicines and misused drugs, suicides, cardiovascular disease, and cycling^10-13^.

During the COVID-19 pandemic, healthcare professionals in England and Wales called for the deaths of their colleagues to be reported to coroners and for PFDs to be issued^14,15^. However, PFDs issued during the COVID-19 pandemic have not been analysed. We therefore aimed to systematically analyse PFDs in which SARS-CoV-2 was directly or indirectly implicated in a death.

## Methods

We extracted a series of cases from the Courts and Tribunals Judiciary website and analysed them according to a study protocol that was preregistered on an open repository^16^.

### Data collection

PFDs are published on the Courts and Tribunals Judiciary website^17^. We used web scraping to systematically collect all published PFDs and created a searchable database, the Preventable Deaths Database^18^, which can be efficiently updated. The code for the web scraper is openly available on GitHub and the methods have been previously described^19,20^. The Preventable Deaths Database contains the case reference number; the date of the report; the name of the deceased; the coroner’s name; the coroner’s jurisdiction; the category of death (as assigned by the Chief Coroner’s office); to whom the report was sent; and the URL to the Judiciary website.

### Eligibility of cases

We screened all 510 PFDs in the Preventable Deaths Database, dated from 01 January 2020 to 28 June 2021, for cases that mentioned COVID-19 as a direct or indirect cause of death. Direct deaths were defined as those that the coroner explicitly attributed to COVID-19 as a cause of death or were associated with a positive test for COVID-19 within 28 days of death. Indirect deaths were defined as those that were not medically caused by COVID-19, but where coroners mentioned that the associated mitigation measures during the COVID-19 pandemic (for example, missed appointments due to lockdown), had contributed to the death. Cases that did not mention SARS-CoV-2 or an effect of the COVID-19 pandemic were excluded.

### Data extraction

For included cases, one study author (BS) manually extracted the following variables into a predesigned Google Sheet, which was cross-examined by another study author (GCR): the individuals or organizations to whom reports were sent and who responded; date of death; age; sex; setting or location of death; medical cause(s) of death; the coroner’s conclusion(s) of the inquest; relevant medical, mental health and social history; substance(s) implicated in the death and the type of substance(s); and the coroner’s concerns. The data available for extraction were limited by the information reported by coroners in the PFDs.

### Data analysis

We used descriptive statistics to describe the numbers and types of cases that met the eligibility criteria for inclusion. The numbers and types of individuals and organizations who received PFDs were synthesized and response rates to coroners were calculated. A response rate of 100% meant that the individual or organization responded to all published PFDs issued by coroners. Once all concerns were extracted, one study author (BS) read through all the concerns and used content analysis to count and classify each concern inductively^21^.

### Software

We used Tableau to present the coroners’ concerns visually and Data Wrapper to geographically map the number of PFDs reported in each region of England and Wales.

### Patient and public Involvement

Patients and the public were not involved in this study.

## Results

Figure 1 shows the results of the extraction process. Of 510 PFDs reported on the Judiciary website between 01 January 2020 and 28 June 2021, 23 (4.5%) were attributed to COVID-19 and deemed preventable by coroners (Table 1).

**Table 1:** Summary of 23 deaths involving SARS-CoV-2 as reported in Prevention of Future Death reports in England and Wales between 1 January 2020 and 28 June 2021, ordered by causes and date of death (created by the authors)

|  |  | Dates |  |  |  | Responses to PFDs |  |
| --- | --- | --- | --- | --- | --- | --- | --- |
| Age | Sex | Death | Inquest | Report | Causes of death | Addressee(s) | Date of reply<sup>±</sup> |
| Directly-related to COVID-19 |  |  |  |  |  |  |  |
| - | M | 13/04/2020 | 14/04/2020 | 01/12/2020 | 1) Community-acquired pneumonia<br>2) COVID-19<br>3) Dementia, chronic obstructive pulmonary disease, asbestos-related pulmonary fibrosis, pleural plaques, type 2 diabetes | 1) CQC<br>2) Vicarage Residential Care Home<br>3) Public Health England<br>4) NHS England<br>5) Greater Manchester Health and Social Care Partnership | 1) 04/02/2021<br>2) Undated<br>3) 26/01/2021<br>4) 02/03/2021<br>5) 19/02/2021 |
| 86 | F | 17/04/2020 | 03/07/2020 | 11/12/2020 | Natural causes and COVID-19 | Whipps Cross Hospital | Received but not dated |
| 74 | M | 21/04/2020 | 22/04/2020 | 01/12/2020 | 1) Hypovolemic shock<br>2) End-stage kidney disease<br>3) Polyneuropathy, frailty and COVID-19 | 1) Department of Health and Social Care<br>2) Royal London Hospital | Not yet received<sup>^</sup> |
| - | M | 21/05/2020 | 26/05/2020 | 09/12/2020 | 1) COVID-19 pneumonia<br>2) Right sided neck of femur fracture, hypertension, atrial fibrillation | 1) Public Health England<br>2) NHS England | 1) 11/02/2021<br>2) 09/02/2021 |
| 18 | M | 31/07/2020 | 03/08/2020 | 30/03/2020 | Drowning to which COVID-19 and asthma were contributory | 1) Craven District Council<br>2) Yorkshire Dales National Park<br>3) Yorkshire Water | Not yet received<sup>^</sup> |
| - | M | 07/09/2020 | 07/09/2020 | 24/04/2021 | 1) Bronchopneumonia in combination with COVID-19<br>2) Falls with vertebral fractures, type 2 diabetes mellitus, pulmonary fibrosis, heart failure and epilepsy | 1) Greater Manchester Health and Social Care Partnership<br>2) NHS England | 1) 07/09/21<br>2) 02/07/21 |
| - | M | 15/11/2020 | 16/11/2020 | 11/06/2021 | 1) Covid 19 Pneumonitis<br>2) Chronic obstructive pulmonary disease, ischaemic heart disease, previous right upper lobe resection for lung adenocarcinoma, type 2 diabetes mellitus | Tameside CCG | 24/06/2021 |
| 90 | M | 28/01/2021 | 08/02/2021 | 23/04/2021 | 1) COVID-19 pneumonia<br>2) Dementia, heart failure, acute on chronic subdural haematoma, fall | Medway Maritime Hospital | 07/06/2021 |
| - | M | 05/02/2021 | 08/02/2021 | 10/06/2021 | 1) COVID-19 on background of immunomodulatory treatment<br>2) Seborrheic eczema<br>3) Peripheral vascular disease | 1) NHS England<br>2) Secretary of State of Health | Not yet received<sup>^</sup> |
| - | M | 03/04/2021 | 17/11/2020 | 14/06/2021 | 1) Aspiration pneumonia on a background of a choking incident, COVID-19 pneumonitis<br>2) Alzheimers dementia | 1) MHRA<br>2) NHS Stockport CCG | 1) 27/07/21<br>2) 06/08/21 |
| - | M | - | 30/10/2020 | 19/02/2021 | COVID-19 pneumonitis | 1) Brighton Sussex University NHS Hospital Trust<br>2) West Sussex NHS Hospital Trust<br>3) Medico-Legal | 1) 19/03/2021<br>2) Not yet received<sup>^</sup><br>3) Not yet received<sup>^</sup> |
| **Indirectly-related to COVID-19** |  |  |  |  |  |  |  |
| 32 | F | 19/03/2020 | 30/03/2020 | 23/11/2020 | 1) Hanging<br>2) Bipolar affective disorder | 1) Sussex Partnership Foundation NHS Trust<br>2) Brighton and Hove City Council | 1) 10/02/2021<br>2) 10/02/2021 |
| - | M | 17/04/2020 | 24/04/2020 | 07/12/2020 | Methadone toxicity | 1) Public Health England<br>2) Haverhill Pharmacy | 1) 13/01/2021<br>2) Undated |
| - | M | 24/04/2020 | 05/05/2020 | 19/11/2020 | Suicide | 1) Woolwich Station Medical Centre<br>2) Ministry of Defence | 1) Not received<sup>^</sup><br>2) 16/02/2021 |
| - | F | 20/06/2020 | 23/06/2020 | 11/02/2021 | 1) Bronchopneumonia<br>2) Frailty<br>3) Dementia<br>4) Hypertension | 1) CQC<br>2) Department of Health and Social Care | 1) 04/06/2021<br>2) 03/06/2021 |
|  |  |  |  |  | 5) Fractured neck of femur |  |  |
| 28 | M | 10/08/2020 | 14/08/2020 | 15/03/2021 | Suicide | Sussex Partnership NHS Foundation Trust | Not yet received<sup>^</sup> |
| 76 | F | 28/09/2020 | 03/12/2020 | 16/12/2020 | Atherosclerosis and complete blockage of one artery | 1) NHS Pathways*<br>2) COVID-19 Pandemic Response Service | 10/02/2021 |
| - | M | 25/10/2020 | 26/10/2020 | 02/06/2021 | Combined drug toxicology | Stockport CCG | 07/07/21 |
| 77 | M | - | 08/10/2020 | 02/02/2021 | 1) Advanced dementia<br>2) Fractured neck of femur<br>3) Ischaemic heart disease | 1) Adult Social Services, Norfolk County Council<br>2) Norfolk and Norwich University Hospital | 1) 11/03/2021<br>2) 09/04/2021 |
| 88 | M | - | 20/08/2020 | 05/02/2021 | 1) Bronchopneumonia<br>2) Heat stroke<br>3) Dehydration | Care Outlook Ltd | 18/04/2021 |
| - | M | - | 11/08/2020 | 07/05/2021 | 1) Small bowel obstruction and perforation<br>2) Ingestion of foreign body | Norfolk and Norwich University Hospital NHS Foundation Trust | 23/07/21 |
| 68 | F | - | 20/09/2020 | 14/12/2020 | 1) Pneumothorax<br>2) Rib fractures<br>3) Fall<br>4) COPD and IHD | West Midlands Ambulance Service | 08/01/2021 |
| 87 | M | - | 08/10/2020 | 07/05/2021 | 1) Urosepsis<br>2) Long term indwelling catheter not changed since October 2019<br>3) Alzheimer's dementia, cerebrovascular accident, chronic kidney disease, bladder cancer, prostate cancer | Lower Clapton Group Practice | 24/06/21 |

\*NHS Digital responded on behalf of NHS Pathways and the COVID-19 Pandemic Response Service. <sup>^</sup>Recipients of PFDs have 56 days from the date of report to respond to the coroner under Regulation 29 of The Coroners (Investigations) Regulations 2013. <sup>^</sup>replies were still overdue on 18/10/21; however not all replies are posted on the Judiciary website.
CCG: Clinical Commissioning Group; CQC: Care Quality Commission; COPD: Chronic Obstructive Pulmonary Disease; MHRA: Medicines and Healthcare products Regulatory Agency; NHS: National Health Service. Deaths directly related were defined as those that the coroner explicitly reported COVID-19 as a cause of death or a positive test for COVID-19 within 28 days of death. Indirectly related deaths were defined as those that were attributed to mitigation measures during the COVID-19 pandemic.

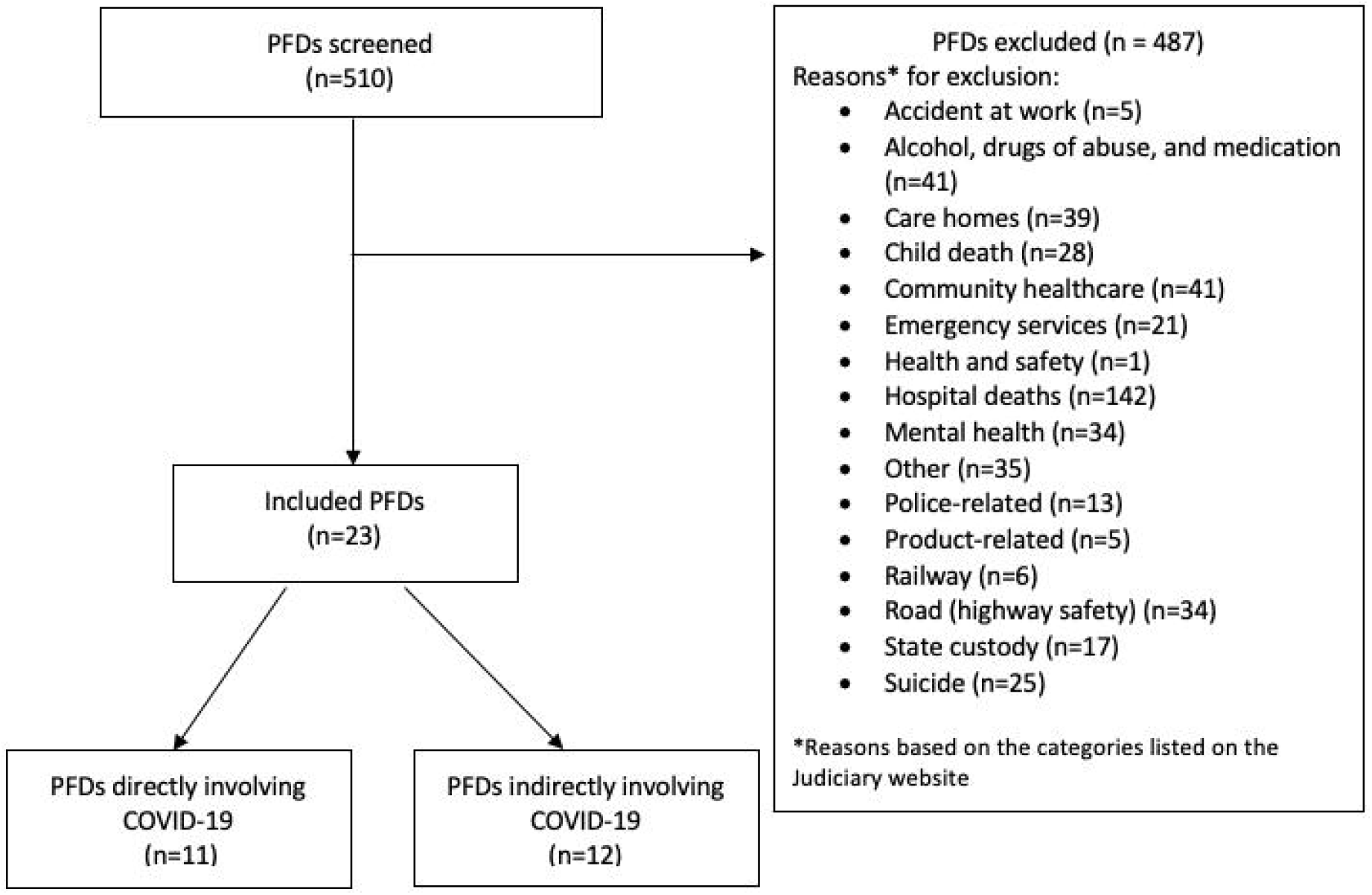

Most of the deaths (78%; n=18) occurred in men, and the median age at death was 76 years (IQR: 50–87 years; range: 18–90 years; n=11). The deaths occurred between 13 April 2020 and 03 April 2021, but the date of death was unreported in six cases.

### Causes of death

Eleven deaths (48%) were directly related to COVID-19, defined as those for which the coroner explicitly reported COVID-19 as a cause of death or that were associated with a positive test for COVID-19 within 28 days of death. Risk factors for death from COVID-19 included type 2 diabetes mellitus, kidney disease, hypertension, heart failure, and epilepsy (Table 1). Pneumonitis co-occurred in 64% of deaths (n=7). One death was attributed to drowning, to which COVID-19 and asthma contributed. In one-third (36%; n=4) of the direct deaths, patients contracted COVID-19 as in-patients for other reasons, and another contracted COVID-19 while a care-home resident. One man’s symptoms of COVID-19 were undiagnosed during a telephone appointment and untreated, resulting in death. In four cases (36%) it was unclear how the deceased had contracted COVID-19.

Twelve deaths (52%) were indirectly related to COVID-19, defined as those that were attributed to mitigation measures during the pandemic. There were three suicides and two cases of inappropriate prescription of medications during lockdown. Two deaths occurred because the deceased refused to go to hospital or a care home, against medical advice, owing to fears of COVID-19. A further two deaths occurred from complications with medical devices when medical appointments were cancelled owing to lockdown and inadequate follow-up; a catheter was not changed in one case, causing urosepsis, and a salivary bypass tube was accidentally left inside the patient, where it perforated the small bowel. In two cases, deaths were attributed to undiagnosed medical problems after incorrect diagnoses of COVID-19 via remote telehealth appointments. One death was due to natural causes, contributed to by several falls and a fractured neck of femur in a patient with advanced dementia.

The 23 deaths were classified into nine groups by the Chief Coroner’s Office; hospital-related (28%; n=11), community healthcare (18%; n=7), care homes (13%; n=5), other (13%; n=5), emergency services (10%; n=4), alcohol, drugs of abuse, and medications (8%; n=3), mental health related (5%; n=2), suicide (3%; n=1), and service personnel related (3%; n=1).

### Geographical variation

Twelve coroners across 12 jurisdictions wrote PFDs relating to COVID-19. Most were written by coroners in the North West of England (39%; n=9), followed by the South East (22%; n=5) and London (17%; n=4) (Figure 2; Supplementary Table 1). Coroners in the South West, North East, West Midlands, and Wales did not report any deaths deemed preventable from COVID-19.

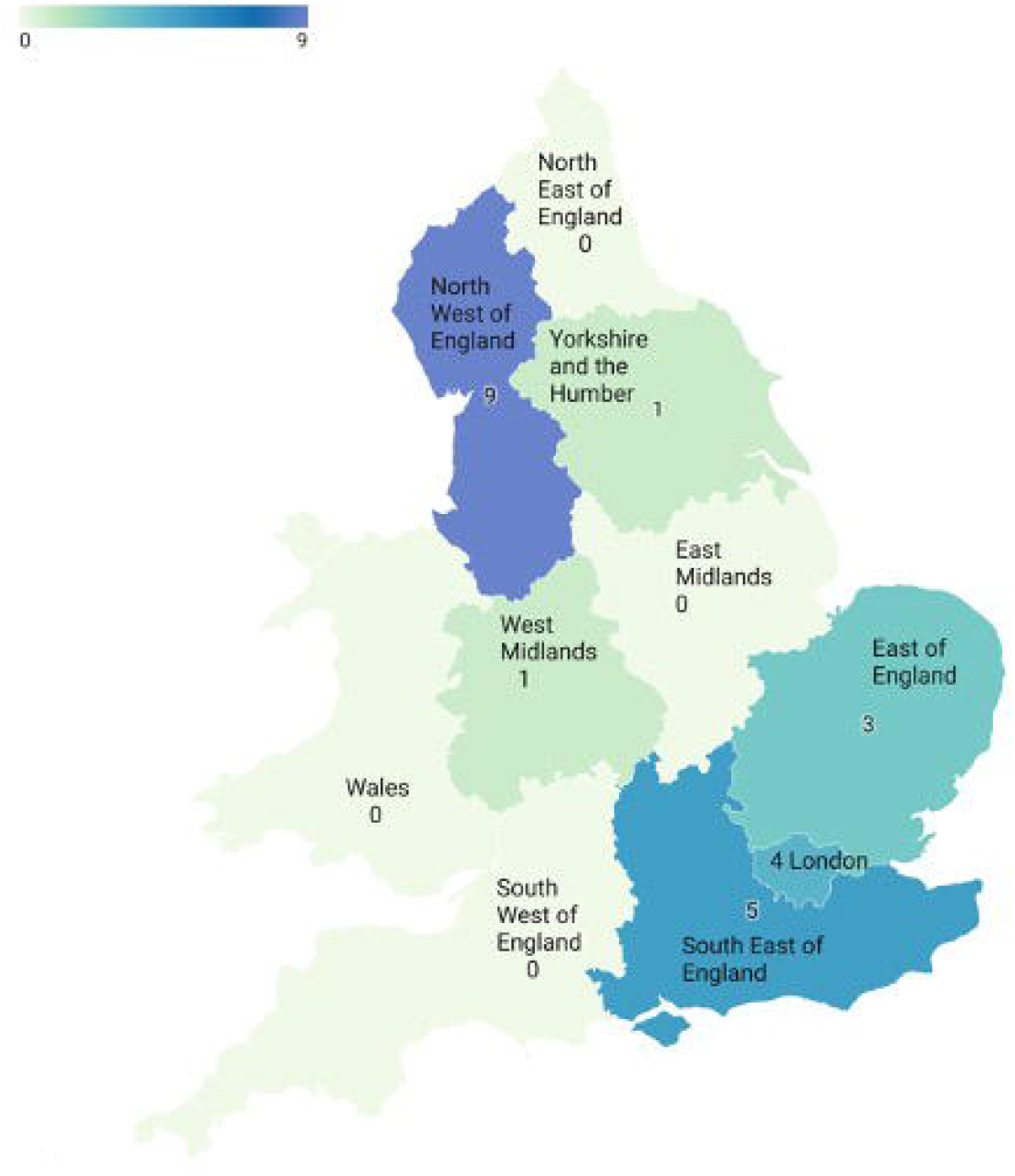

### Coroners’ concerns

The coroners raised 56 individual concerns in the 23 PFDs. We categorized them into 28 groups and five higher-order categories (Figure 3; Supplementary Table 2). Poor communication was reported in one-third of PFDs, followed by failure to follow protocols (23%), lack of education and training (19%), lack of resources (16%), and safety concerns (12%).

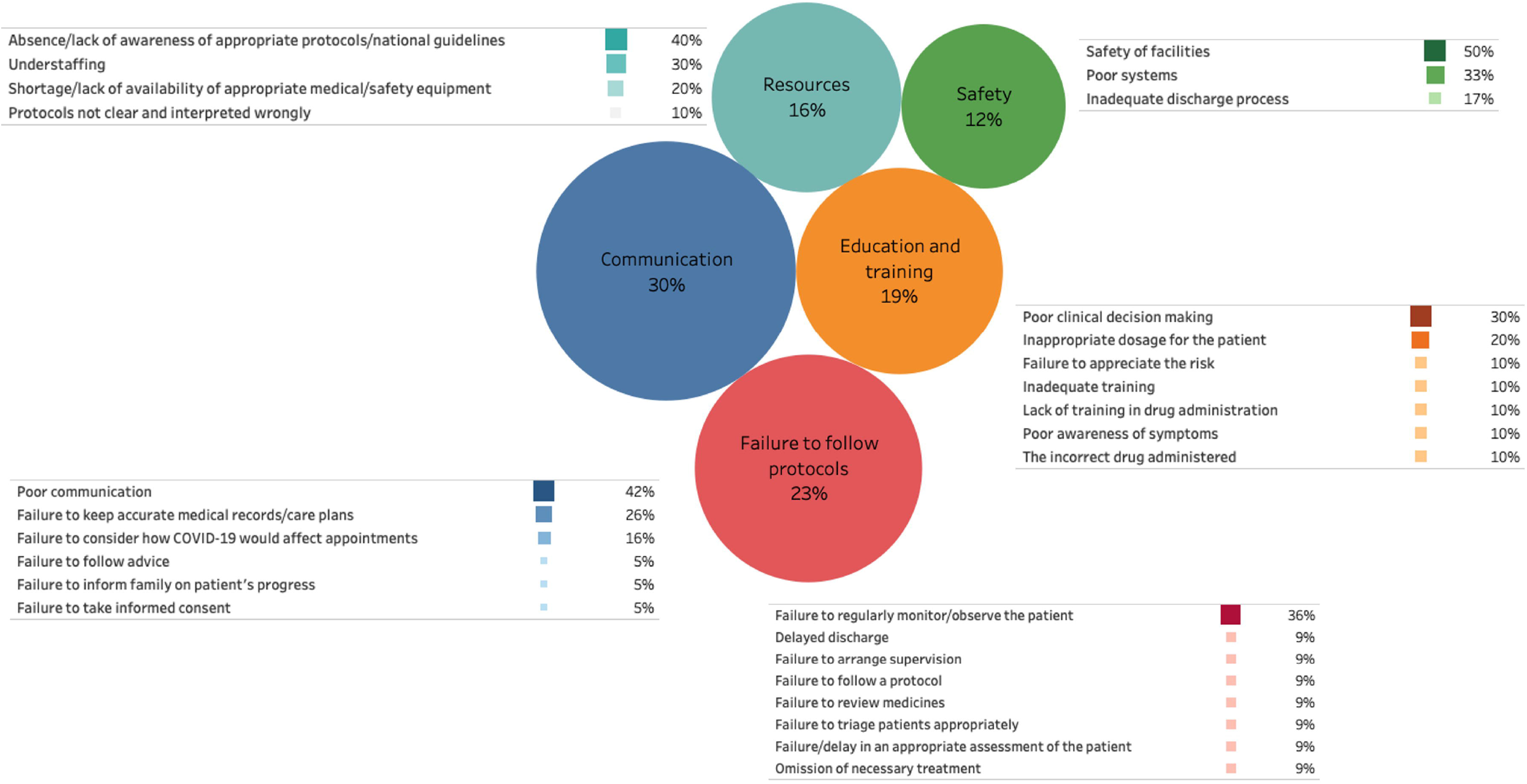

#### 1) Communication

Most (42%, n=8) concerns involved poor communication, followed by failure to keep accurate medical records/care plans (n=5) and failure to consider how the COVID-19 pandemic would affect appointments (n=3).

**Example 1:** *An elderly woman residing in a care home, at high risk of falls, became unwell. Her GP remotely diagnosed suspected COVID-19, and she was isolated in her room, with staff observation and sensor mats to ensure her wellbeing. She fell while unobserved and was admitted to hospital with a fractured neck of femur, bronchopneumonia, and possible COVID-19. A test for SARS-COV-2 was negative. She was unfit for surgery, deteriorated, and later died in hospital from bronchopneumonia. The care home had a risk plan that stipulated that she was to be observed during the day in communal areas. However, the home was not staffed to provide one-to-one observation for residents in self-isolation*.

The coroner believed that it was unclear how care homes were being advised to safely manage residents at risk of falls when isolation was required, and the home knew of no guidance they could follow to manage the risk. Furthermore, restrictions on family visitors when in hospital presented significant communication problems, which reduced appropriate support and timely clinical decision making.

#### 2) Failure to follow protocols

The failure to regularly monitor/observe the patient was the most common form of failure (36%; n=4), followed by a general failure to follow protocols, omission of necessary treatment, and delayed discharge (Figure 3).

**Example 2:** *A 28-year-old man with a lifelong history of low mood and depression with suicidal ideation was socially isolated, which was exacerbated when he was furloughed from his job. He had made five or six attempts to kill himself in 2020 alone and came to the attention of an NHS Trust in May 2020. He registered with a GP at around the same time and was assessed by a Mental Health practitioner, who referred him for treatment. The man was aware of the referral and waited for treatment, but unfortunately the referral was lost. He died by suicide at home 2 months later*.

The coroner concluded that the Care Programme Approach (CPA), as set out in national guidance “Refocusing the CPA – Policy and Positive Practical Guidance”, had not been followed. It was clear at the inquest that staff members were not aware of how matters should be dealt with and that this depended heavily on geographical location.

#### 3) Education and training

The most common concern was poor clinical decision making (30%, n=3), followed by inappropriate dosage of medication (20%, n=2).

**Example 3:** *A man with drug dependency had been receiving support from a Recovery Network and was generally fit, well, and in good spirits. He received a regular prescription for methadone in daily dosage bottles three times a week. During the COVID-19 pandemic this regimen was changed to once every 14 days, giving him access to a much larger quantity of methadone. The deceased was found at home with a very high blood concentration of methadone. There was no evidence that he intended to take his own life. At the start of the pandemic, Public Health England (PHE) guidance was issued that patients taking short-term methadone should be moved to long-term treatment. The doctor who changed the prescription stipulated that the drug must be supplied in single daily dosage bottles, and explanatory letters had been sent to all the pharmacies that supplied opiate replacement therapies to his patients*.

The coroner saw evidence that the prescription was not in daily dose bottles and that there was no measuring jug to enable accurate measurement of doses. The coroner believed that it was probable that the man had guessed his first dose from the large methadone bottle. If he had been given his daily dosage bottles, as prescribed, a measuring jug, and instructions on how to use it, his death might have been prevented.

#### 4) Resources

Lack of awareness of appropriate protocols and national guidelines was the most common failure (40%, n=4), followed by understaffing (30%, n=3), a shortage or lack of availability of medical equipment (20%, n=2), and unclear protocols (10%, n=1).

**Example 4:** *A man was admitted to hospital after an accidental fall at home. After surgery for a fractured hip, he developed a chest infection. When fit for discharge, he was moved to several different wards and eventually put in a bay where patients had been exposed to COVID-19. He subsequently tested positive for COVID-19, deteriorated rapidly, and died*.

The coroner heard that the decision to move the deceased had been made on interpretation of guidance from PHE. The Trust changed its policy, and such movements are reportedly no longer taking place. However, when the PFD was written the guidance from PHE had not been amended, and it was not known how other Trusts were choosing to interpret the guidance, potentially putting other vulnerable in-patients at risk of COVID-19. Rapid and national dissemination of coroner’s concerns might have prevented similar errors in other hospitals.

#### 5) Safety

Half of the concerns about safety related to facilities (50%, n=3), followed by poor systems (33%, n=2) and inadequate discharge processes (17%, n=1).

**Example 5:** *A resident in a care home, who had pulmonary fibrosis, had an unwitnessed fall. He lay on the floor for over 4 hours awaiting an ambulance. Sepsis was diagnosed and it was thought that he had symptoms consistent with COVID-19. He remained at the home until 11 April, became unresponsive, deteriorated rapidly, was moved to palliative care, and died on 13 April 2020*.

The coroner reported that the care had been of limited quality, notwithstanding the diagnosis of COVID-19 and his vulnerability. The care home was unclear if staff had brought COVID-19 into the home or if admission of residents from the community who had not being tested before admission had caused COVID-19 to enter the home. There was no risk assessment in place relating to the admission of new residents, creating an unsafe environment for vulnerable people.

### Responses to PFDs

Forty-three unique individuals and organizations received PFDs from coroners (Table 2). Most reports were sent to the NHS (51%; n=22), followed by the Government and related bodies, other organizations, and professional bodies. Government organizations had the highest response rates (64%; n=7), followed by professional bodies (50%; n=2), other organizations (50%; n=3), and NHS organizations (45%; n=10). Of the 23 PFDs, nine had a 100% response rate and ten had no responses on the Judiciary website.

**Table 2:** Recipients of Prevention of Future Deaths reports involving COVID-19 in England and Wales between 1 January 2020 and 28 June 2021 and their response rates (created by the authors)

| Addressee | No. of PFDs sent | No. of responses<sup>±</sup> | Response rate (%) |
| --- | --- | --- | --- |
| **NHS organizations** | **22** | **10** | **45%** |
| Trusts | 5 | 2 | 40% |
| NHS England | 4 | 2 | 50% |
| NHS Hospitals | 4 | 3 | 75% |
| CCGs | 3 | 0 | 0% |
| Health and Social Care Partnerships | 2 | 1 | 50% |
| NHS Pathways* | 1 | 1 | 100% |
| Ambulance services | 1 | 1 | 100% |
| GPs | 2 | 0 | 0% |
| **Government** | **11** | **7** | **64%** |
| Public Health England | 3 | 3 | 100% |
| Department of Health and Social Care | 2 | 1 | 50% |
| Local authorities | 3 | 2 | 67% |
| COVID-19 pandemic response service* | 1 | 1 | 100% |
| Secretary of State of Health | 1 | 0 | 0% |
| Ministry of Defence | 1 | 0 | 0% |
| **Professional bodies** | **4** | **2** | **50%** |
| CQC | 2 | 2 | 100% |
| General Pharmaceutical Council | 1 | 0 | 0% |
| MHRA | 1 | 0 | 0% |
| **Other** | **6** | **3** | **50%** |
| Care homes/providers | 2 | 2 | 100% |
| Water board | 1 | 0 | 0% |
| National Park | 1 | 0 | 0% |
| Legal | 1 | 0 | 0% |
| Pharmacy | 1 | 1 | 100% |

\*NHS Digital responded on behalf of NHS Pathways and the COVID-19 Pandemic Response Service
‡Recipients of PFDs have 56 days from the date of the report to respond to the coroner under Regulation 29 of The Coroners (Investigations) Regulations 2013
CCG: Clinical Commissioning Group; CQC: Care Quality Commission; GPs: General Practitioners; MHRA: Medicines and Healthcare products Regulatory Agency; NHS: National Health Service

## Discussion

One in 20 PFDs published online involved COVID-19. Most of the deaths occurred in men and older adults. There was wide geographical variation: no PFDs were reported by coroners in Wales or in the North East, East Midlands, or South West of England. Coroners raised several concerns, particularly regarding problems with communication and following protocols. The largest numbers of PFDs were sent to NHS Trusts and the Government.

Eleven deaths were directly caused by SARS-CoV-2. In one-third of these cases, patients acquired COVID-19 after admission to hospital for an unrelated reason, and another acquired COVID-19 while in a care home. This suggests that measures to reduce the transmission of SARS-COV-2 in healthcare settings were not adequate. Healthcare settings should focus on evidence-based provisions, such as ventilation, personal protective equipment (PPE), and regular testing to mitigate the risk to vulnerable patients^22^.

Twelve deaths were indirectly attributed to the COVID-19 pandemic. This finding highlights the importance of considering the harms of measures and policies that were implemented to reduce the transmission of SARS-COV-2 in the community. Reduced social interactions and changed working conditions or loss of work and income have negatively affected adult mental health in the UK^23^. Six million patients in the UK did not seek treatment in 2020 (so-called “missing patients”), owing to reprioritization of healthcare services^24^. In some cases reduced access to care because of lockdown, despite telemedicine, led to deaths.

Healthcare professionals in England and Wales have called for the deaths of their colleagues to be reported to coroners and for PFDs to be issued^14,15^. However, we did not identify any PFDs that reported deaths of healthcare professionals.

The UK Government has stated that they will begin their public inquiry into the handling of the COVID-19 pandemic to learn lessons for future pandemics^25^. We have identified several areas that the Government should address during this inquiry, including poor communication and gaps in education and training. PFDs should be examined by the Government and healthcare providers to inform quality improvement and patient safety initiatives.

The Office of the Chief Coroner, who is responsible for uploading PFDs to the Courts and Tribunals Judiciary website, categorized the COVID-19-related PFDs into nine groups, including hospital-related, community healthcare, and care homes. A new category specifically for the effects of pandemics should be added to the Judiciary website to assist the Government in examining these case reports so that policy measures can be implemented for future pandemics.

Our study has several limitations. The 23 deaths do not represent all deaths during the COVID-19 pandemic that could have been prevented in England and Wales. We are also limited by the information reported by coroners in PFDs. In 52% of PFDs the age of the deceased was not reported and 26% did not report the date of death. There were also regions in England and Wales that did not report any PFDs.

Under-reporting of PFDs limits the capacity for actions to be taken to prevent future deaths. However, it is likely that more PFDs relating to COVID-19 will be published, owing to the backlog of inquests and the time it takes for inquests to conclude and PFDs to be written and published.

There are no clear guidelines in England and Wales for referring deaths to coroners nor for determining when a PFD should be issued and what information to include, hence the missing data. There is also no auditing or quality control of PFDs and their responses. Thus, whether action is taken to prevent such deaths, and the timeliness of such action, is unknown and unmonitored. In the meantime, we encourage coroners across England and Wales to continue writing PFDs when they believe that deaths could have been prevented.

PFDs contain a rich source of information that can be systematically analysed to share information on preventing harms.The concerns identified in the 23 PFDs should be considered during the UK Government’s inquiry, including how communication, protocols, education, training, resources, and patient safety can be improved. Guidelines on how and when to report deaths to coroners, thresholds for issuing PFDs, and the necessarymandatory types of information to report in PFDs, including age and date of death, are needed. Coroners in England and Wales should be encouraged to continue writing PFDs during the COVID-19 pandemic, particularly when deaths involve frontline healthcare professionals.

## Data Availability

The data, statistical code, and study materials are openly available via the Open Science Framework (OSF) and GitHub
Richards, G. georgiarichards.github.io. (Github).
Swift, B., Heneghan, C., Aronson, J. K., Howard, D. & Richards, G. C. Preventable deaths from SARS-CoV-2 in England and Wales. OSF https://osf.io/bfypc/ (2021) doi:10.17605/OSF.IO/BFYPC.

## Declarations

### Statement of ethical approval

We are using publicly available information, for which ethics committee approval is not required.

### Contributorship statement

GCR developed the idea for this study, wrote the initial study protocol, ran the python code to collect the most recent PFDs for screening, contributed to the first draft of the manuscript, and provided supervisory support. BS contributed to the study protocol, screened the 510 PFDs for their eligibility, extracted the data from the 23 included cases, analysed the data, and wrote the first draft of the manuscript. CH, JKA, and DJH contributed to the study protocol, data analysis, and supervision of the project. All authors read, reviewed, and approved the manuscript before submission.

### Data sharing

The data, statistical code, and study materials are openly available via the Open Science Framework (OSF) and GitHub^16,19^.

