## Supplementary material for "Preventable deaths from SARS-CoV-2 in England and Wales: a systematic case series of coroners’ reports during the COVID-19 pandemic"

| **Region** | **No. of cases (%)** |
| --- | --- |
| **North West England** | **9 (39.1)** |
| Greater Manchester | 6 (26.1) |
| Manchester South | 2 (8.7) |
| County of Cumbria | 1 (4.3) |
| **South East** | **5 (21.6)** |
| Brighton and Hove | 3 (13.0) |
| Mid Kent and Medway | 1 (4.3) |
| North East Kent | 1 (4.3) |
| **London** | **4 (17.3)** |
| Inner North London | 3 (13.0) |
| East London | 1 (4.3) |
| **East of England** | **3 (13.0)** |
| Norfolk | 2 (8.7) |
| Suffolk | 1 (4.3) |
| **Yorkshire and the Humber** | **1 (4.3)** |
| North Yorkshire | 1 (4.3) |
| **West Midlands** | **1 (4.3)** |
| Black County | 1 (4.3) |
| **North East** | 0 |
| **East Midlands** | 0 |
| **South West** | 0 |
| **Wales** | 0 |

**APPENDIX**

**Supplementary table 1**: Jurisdictions of coroners who issued Prevention of Future Deaths reports involving SARS-COV-2 in England and Wales between 01 January 2020 and 28 June 2021. Created by the authors.

**Supplementary table 2**: Concerns raised by coroners in Prevention of Future Deaths reports involving COVID-19  in England and Wales between 01 January 2020 and 28 June 2021. Created by the authors.

| **Concerns** | **No. of cases (%)** |
| --- | --- |
| **Communication (30%)** | |
| Poor communication | 8 (42) |
| Failure to keep accurate medical records/care plans | 5 (26) |
| Failure to consider how COVID-19 would affect appointments | 3 (16) |
| Failure to take informed consent | 1 (5) |
| Failure to follow advice | 1 (5) |
| Failure to inform family on patient's progress | 1 (5) |
| **Failure to follow protocols (23%)** | |
| Failure to regularly monitor/observe the patient | 4 (36) |
| Failure to follow a protocol | 1 (9) |
| Omission of necessary treatment | 1 (9) |
| Delayed discharge | 1 (9) |
| Failure/delay in an appropriate assessment of the patient | 1 (9) |
| Failure to triage patients appropriately | 1 (9) |
| Failure to arrange supervision | 1 (9) |
| Failure to review medicines | 1 (9) |
| **Resources (16%)** | |
| Absence/lack of awareness of appropriate protocols/national guidelines | 4 (40) |
| Understaffing | 3 (30) |
| Shortage/lack of availability of appropriate medical/safety equipment | 2 (20) |
| Protocols not clear and interpreted wrongly | 1 (1) |
| **Education and training (19%)** | |
| Poor clinical decision making | 3 (30) |
| Inappropriate dosage for the patient | 2 (20) |
| Inadequate training | 1 (10) |
| Poor awareness of symptoms | 1 (10) |
| Failure to appreciate the risk | 1 (10) |
| Lack of training in drug administration | 1 (10) |
| The incorrect drug administered | 1 (10) |
| **Safety (12%)** | |
| Safety of facilities | 3 (50) |
| Poor systems | 2 (33) |
| Inadequate discharge process | 1 (17) |
